# Comparing the clinical trial characteristics of industry-funded trials and non–industry-funded trials

**DOI:** 10.1101/2023.03.24.23287707

**Authors:** Emily Hughes, Tamara Van Bakel, Ashley Raudanskis, Prachi Ray, Benazir Hodzic-Santor, Ushma Purohit, Chana A. Sacks, Michael Fralick

**Affiliations:** Sinai Health System, Division of General Internal Medicine, Toronto, Ontario, Canada; Lunenfeld-Tanenbaum Research Institute, Sinai Health System, Toronto, Ontario, Canada; Division of General Internal Medicine and Mongan Institute, Massachusetts General Hospital, Harvard Medical School, Boston, USA

**Keywords:** randomized controlled trials, funder, trial characteristics, pharmaceutical, industry, natural language processing

## Abstract

**Importance:** In randomized controlled trials (RCTs), small sample size and lack of blinding can cause biased and spurious results. Whether and how study characteristics differ based on a trial’s funder is an important area to study.

**Objective:** To compare study characteristics of RCTs funded by industry with study characteristics of RCTs not funded by industry.

**Design, Setting and Participants:** We systematically reviewed all RCTs published between 2015 and 2019 in the New England Journal of Medicine (NEJM), Lancet, and Journal of the American Medical Association (JAMA). Our primary data sources were ClinicalTrials.gov and MEDLINE. Data extraction included manual review and use of natural language processing.

**Main Outcomes and Measures:** We compared the rate of blinding, use of placebo, and sample size. We used natural language processing to analyze the sentiment of the study’s conclusion as reported in the abstract. As proxies for knowledge dissemination, we calculated the AltMetric scores and number of times the article was cited (citation count).

**Results:** We identified 1533 RCTs published by NEJM, Lancet, and JAMA between 2015 and 2019. Of these RCTs, 697 were funded by industry. Trials funded by industry were more likely to be blinded (n=378, 54% vs n=318, 38%), more likely to include a placebo (n=317, 45% vs n=196, 23%), more likely to post their results on ClinicalTrials.gov (78%, 443 of 570 vs 41%, 207 of 501) compared to trials that were not industry funded. Industry-funded RCTs had a smaller sample size than non–industry-funded RCTs (median=557 [IQR: 230, 1369] vs 648 [IQR: 301, 1916], P<0.01). Trials funded by industry had more citations than non–industry-funded trials (285 vs 145, p < 0.01), while per-manuscript Almetric scores were similar between both groups (229 vs 226, p=0.2).

**Conclusions and Relevance:** These data highlight important variability in key metrics of trial quality and call attention to specific areas of improvement, especially for non–industry-funded trials.

## INTRODUCTION

In 1948, the Medical Research Council’s trial of streptomycin for the treatment of pulmonary tuberculosis became the first published randomized controlled trial (RCT) in medicine.^1^ The RCT design is now considered to be the gold standard for generating rigorous and reliable data to inform clinical decision making.^2^ Randomization removes selection bias and, assuming the trial is sufficiently large, balances measured and unmeasured covariates.^3,4^ Randomization can also ensure that patients have a consistently defined study start date, which prevents immortal time bias.^5^ When investigators conducting RCTs blind participants, clinicians, and researchers, the risk of ascertainment bias and observer bias are minimized, which helps to prevent time-varying confounding.^3^ Maintaining the rigor of a well-designed and well-conducted RCT requires considerable time and financial investment. The average cost of an RCT varies depending on the disease area, but in the United States the cost of Phase 3 RCTs range, on average, from $12 million to $53 million.^6^ These costs are often prohibitive for academic investigators.

The potential for bias in industry-sponsored trials has been well documented, and has led to important questions about whether corporate entities with a financial interest in the outcome of trials should be so closely involved in the research.^7^ Concerns have been raised that industry-sponsored trials may be terminated early due to financial reasons rather than scientific or ethical reasons, and that academic investigators who receive corporate funding may be incentivized to bias the analysis and reporting of trial results.^7^ Further, there have been reports of industry choosing to selectively present positive results and withhold negative results. Among physicians, these documented instances have fostered mistrust in industry-sponsored trials.^8^ One well-cited example is GlaxoSmithKline’s marketing of Paxil (paroxetine) for adolescent major depression.

The initial study publication in 2001 indicated that paroxetine for treatment of adolescent major depression was generally well tolerated, and demonstrated significantly greater improvement compared with placebo.^9^ However, a 2015 post-publication analysis of the original raw data revealed that GlaxoSmithKline had manipulated the data and selectively downplayed the harms of the drug: in fact, paroxetine is no better than placebo for any prespecified primary or secondary efficacy outcome, and use could result in serious side effects including self-injury and suicide.^10^

There is consequently—and understandably—a perception among clinicians that RCTs funded by industry may be of lower quality. In a 2012 study, Kesselheim et al.^11^ created a series of abstracts for hypothetical clinical trials and randomly assigned to each the designation of either pharmaceutical company funding, federal funding, or no funding. Board-certified internists were then asked to evaluate the quality of the hypothetical trials. Despite the content of the abstract being otherwise identical, physicians who received the version indicating funding by a pharmaceutical company perceived the methodologic quality more negatively than physicians evaluating the exact same abstract with an indication of either federal funding or no funding.

Systematic analysis of how trial characteristics differ between industry-funded and non–industry-funded trials may offer insight into the extent to which pre-conceived notions of study quality based on funder are accurate. Our objective was to compare study characteristics of RCTs funded by industry to those of RCTs not funded by industry.

## METHODS

### Study Population

We reviewed all RCTs published in the *New England Journal of Medicine* (*NEJM*), the *Journal of the American Medical Association* (*JAMA*), and *Lancet* between January 1, 2015 and December 31, 2019. These journals were selected because they have the highest impact factors among general medicine peer-reviewed journals, and they commonly publish RCTs. We chose to stop data collection at the year 2019 (inclusive) because the advent of the COVID-19 pandemic may have changed the landscape of RCT publishing. Excluding trials after 2019 also afforded at least two years of follow-up in the evaluation of post-publication metrics for each RCT. Using MEDLINE, we identified all articles published in these journals during the study period and then excluded review articles, research letters, letters to the editors, and editorials. The title and abstract of the remaining articles were reviewed independently by three study members (MF, UP, AR) to identify and exclude any remaining non-randomized studies and duplicate publications. Disagreements were resolved through consensus. A detailed outline of the study selection process is provided in Figure 1.

**Figure 1.**
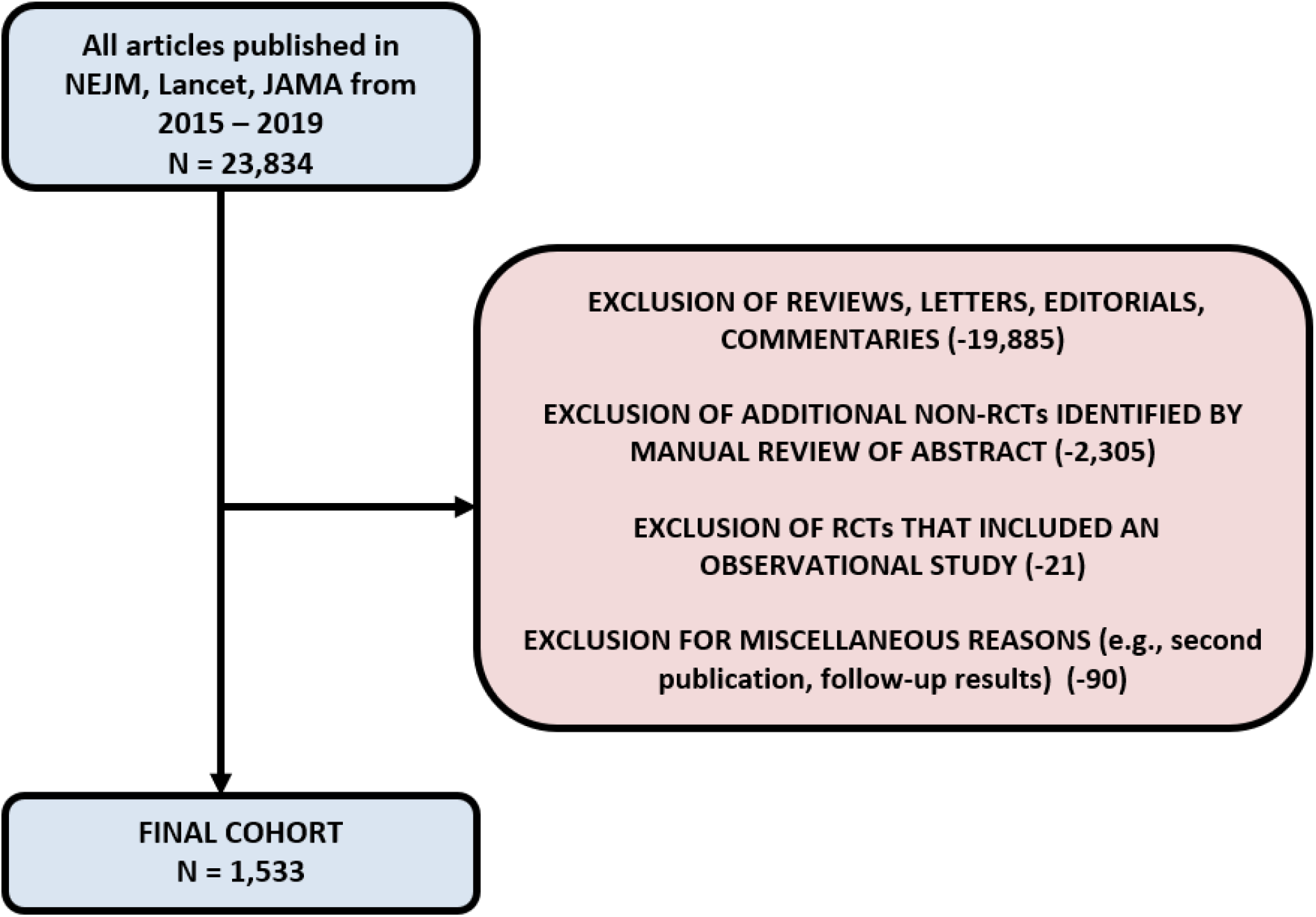
Study Flow Diagram NEJM = New England Journal of Medicine, JAMA = Journal of the American Medical Association, RCT = randomized controlled trial

### RCT Data Collection

For each RCT, data were manually extracted from the study abstract. These datapoints included sample size, blinding, comparator type (i.e., placebo, active comparator, other), disease area, outcome type (i.e., surrogate outcome or not^12^), the study’s conclusion as written in the abstract, the primary outcome result (i.e., positive, neutral, or negative), and funder type. Funder type was determined through manual review of the manuscript and classified as one of “industry-funded,” “non–industry-funded,” or “combination funded.” Combination funded was defined as trials that included both industry and non-industry funders. We then combined “industry-funded” and “combination funded” into one group, classifying the resultant merged group as “industry-funded.” When pharmaceutical companies donated the trial intervention (e.g., drug, device, placebo) but gave no monetary funding, the RCT was considered non–industry-funded. The study outcome was considered positive if the point estimate for the primary outcome identified a benefit and the 95% confidence intervals excluded the null. The study outcome was considered negative if the point estimate for the primary outcome was below the null and the 95% confidence intervals excluded the null. The study outcome was considered neutral if the point estimate included the null. For non-inferiority trials, the study outcome was considered positive if the study was superior or non-inferior. To determine whether results were posted online on ClinicalTrials.gov, we used the site’s API.

### Knowledge Dissemination Data Collection

Altmetric scores were obtained for each RCT. Almetric scores quantify an aggregate of an article’s online attention by including data on volume of mentions, author of online mention (e.g., reporter), and mention source (e.g., Twitter, blog, news report). Citation counts were obtained for each RCT and used as a further measure of knowledge dissemination. Altmetric data, including Altmetric Scores and their breakdown, were obtained using the Altmetric Details Page API via the rAltmetric R package. The number of citations was obtained through the CrossRef API via the rcrossref R package. Using the Digital Object Identifier (DOI) of the peer-reviewed article, data were merged from various sources.

### Study Outcomes & Statistical Analysis

We compared the frequency of double blinding, inclusion of placebo, sample size, and posting of trial results on ClinicalTrials.gov between industry-funded trials and non–industry-funded trials. We also analyzed the sentiment of the study’s conclusion using two different transformer-based models (a neural network-based technique for natural language processing). The models we used have been pre-trained on a large corpus of English text in a self-supervised fashion and routinely applied in sentiment analysis.^13^ Models 1 and 2 both used BERT, a bidirectional transformer model pre-trained using masked language modeling and next sentence prediction.^14^ Model 1 classified responses into either positive or negative, while Model 2 indicated the sentiment of the response as a number of stars (between 1 and 5, where 1 is highly negative and 5 is highly positive). Descriptive statistics were used to compare industry-funded RCTs to non–industry-funded RCTs. All analyses were conducted using R version 3.1.2.5.

## RESULTS

We identified 1533 RCTs published in *NEJM, JAMA* and *Lancet* between January 1, 2015, and December 31, 2019. Most of the RCTs were trials of medications. Approximately one-third were double-blind, approximately 40% had a surrogate outcome as their primary endpoint, and approximately two-thirds had a positive primary outcome (Table 1). Of all the RCTs, 697 were funded by industry while 835 were not; one study lacked funding details altogether.

**Table 1.** Baseline characteristics

|  | Overall<br>N = 1533 | Industry-funded<br>N = 697 | Non–industry-funded<br>N = 835 | SMD (p) |
| --- | --- | --- | --- | --- |
| **Subspecialty Focus** |  |  |  | 0.509 (<0.001) |
| Cardiovascular | 302 (20.1) | 173 (24.8) | 129 (16.0) |  |
| Endocrinology | 67 (4.5) | 32 (4.6) | 35 (4.3) |  |
| Infectious Disease | 182 (12.1) | 55 (7.9) | 127 (15.8) |  |
| Neurology | 100 (6.7) | 46 (6.6) | 54 (6.7) |  |
| Oncology | 220 (14.6) | 144 (20.7) | 76 (9.4) |  |
| Psychiatry | 40 (2.7) | 7 (1.0) | 33 (4.1) |  |
| Respirology | 104 (6.9) | 51 (7.3) | 53 (6.6) |  |
| Other | 488 (32.5) | 189 (27.1) | 298 (37.0) |  |
| NA | 30 (2.0) | 0 (0.0) | 30 (3.6) |  |
| **Published Year** |  |  |  | 0.353 (0.108) |
| 2015 | 318 (20.7%) | 137 (19.66%) | 181 (21.7%) |  |
| 2016 | 277 (18.1%) | 120 (17.2%) | 157 (18.8%) |  |
| 2017 | 292 (19.0%) | 143 (20.5%) | 148 (17.7%) |  |
| 2018 | 280 (18.3%) | 121 (17.4%) | 159 (19.0%) |  |
| 2019 | 366 (23.9%) | 176 (25.3%) | 190 (22.9%) |  |
| **Journal** |  |  |  | 0.357 (<0.001) |
| JAMA | 355 (23.1) | 106 (15.2) | 249 (29.7) |  |
| Lancet | 542 (35.4) | 263 (37.7) | 279 (33.4) |  |
| NEJM | 637 (41.6) | 328 (47.1) | 308 (36.9) |  |
| **Intervention** |  |  |  | 0.744 (<0.001) |
| Drug | 988 (64.4%) | 567 (81.3%) | 421 (50.4%) |  |
| Device/Procedure | 306 (20.0%) | 99 (14.2%) | 206 (24.7%) |  |
| Other | 239 (15.6%) | 31 (4.4%) | 208 (24.9%) |  |
| **Blinding** |  |  |  | 0.494 (<0.001) |
| Double | 537 (35.0%) | 330 (47.3%) | 207 (24.8%) |  |
| Single | 159 (10.4%) | 48 (6.89%) | 111 (13.3%) |  |
| Unblinded | 837 (54.6%) | 319 (45.8%) | 517 (61.9%) |  |
| **Surrogate Outcome** |  |  |  | 0.256 (<0.001) |
| Yes | 638 (41.6%) | 338 (48.5%) | 300 (35.9%) |  |
| **Overall Outcome** |  |  |  | 0.636 (<0.001) |
| negative | 511 (33.3%) | 125 (17.9%) | 386 (46.2%) |  |
| positive | 1022 (66.7%) | 572 (82.1%) | 449 (53.8%) |  |
| **Comparator\*** |  |  |  | 0.681 (<0.001) |
| Active comparator | 678 (44.2%) | 316 (45.3%) | 361 (43.2%) |  |
| Placebo | 513 (33.5%) | 317 (45.5%) | 196 (23.5%) |  |
| **Sample size** |  |  |  | 0.143 (0.007) |
| Median | 619 (IQR:<br>270,1741) | 557.0 (IQR:<br>229.5, 1369.0) | 648 (IQR: 301, 1916) |  |

Legend: SMD = standardized mean difference; JAMA = Journal of the American Medical Association; NEJM = New England Journal of Medicine; IQR = Interquartile range. \*Remaining studies had a comparator that was neither active nor placebo.

Studies funded by industry were more likely to be related to cardiovascular disease or oncology, and less likely to be related to infectious disease (Table 1). The median sample size was 557 (IQR: 230-1369) for industry-funded trials and 648 (IQR: 301, 1916) for non–industry-funded trials. Compared to non–industry-funded trials, industry-funded trials were more likely to be double-blind, more likely to include a placebo, and more likely to have a surrogate primary outcome (Table 1). The primary outcome was positive for 82% of industry-funded RCTs compared to 54% of non–industry-funded RCTs.

We found that the concluding statements of trials funded by industry were more likely to have greater positive sentiment than those not funded by industry (n=246, 35% vs n=208, 25%), which aligns with the fact that industry-funded trials were more likely to have positive findings. Our findings were robust across two different natural language processing (NLP) methods (Table 2).

**Table 2.** Primary outcome of the study compared to the sentiment of the abstract

|  | **Industry-funded**<br>**N=697** | **Non-industry-funded**<br>**N=835** |
| --- | --- | --- |
| **Study's Overall Outcome** |  |  |
| Positive | 572 (82.1%) | 449 (53.8%) |
| Negative | 125 (17.9%) | 386 (46.2%) |
| **Conclusion sentiment using NLP (%)** |  |  |
| Positive | 246 (35.29%) | 208 (24.91%) |
| Negative | 451 (64.71%) | 627 (75.09%) |
| **Conclusion sentiment using NLP 5 star (%)** |  |  |
| 1 star [highly negative] | 16 (2.3%) | 16 (1.92%) |
| 2 stars | 145 (20.8%) | 259 (31.02%) |
| 3 stars | 183 (26.26%) | 268 (32.1%) |
| 4 stars | 281 (40.32%) | 251 (30.06%) |
| 5 stars [highly positive] | 72 (10.33%) | 41 (4.91%) |

Legend: NLP = natural language processing

Industry-funded studies received twice as many citations as non–industry-funded studies (n=286 [IQR: 140-562] vs n=145 [IQR: 76-266], p < 0.01). AltMetric scores were similar for all included studies, regardless of their funding source (n=229 [IQR: 122-468] for industry-funded; n=226 [IQR: 119-441] for non–industry-funded, p=0.2). Industry-funded studies were also more likely to post their results on ClinicalTrials.gov compared to non–industry-funded studies (78% vs 41%).

## DISCUSSION

In this study of RCTs published in *NEJM, JAMA*, and *Lancet* between 2015 and 2019, we identified 697 RCTs that were funded by industry and 835 that were not. Trials funded by industry were more likely to be blinded, more likely to be placebo controlled, and more likely to post trial results on ClinicalTrials.gov. These data suggest that there are key metrics of trial quality that may be better conducted in industry-funded trials compared to non–industry-funded trials, adding important and often overlooked nuance to the broader discussion about industry-funded research.

The greatest strengths of RCTs are that selection bias is removed and, assuming the study is large enough, both measured and unmeasured confounders are balanced. However, without blinding it is almost impossible to prevent time varying confounding or ascertainment bias, both of which can directly affect the internal validity of a study, especially when the primary outcome is subjective or prone to measurement error. Trials funded by industry may have been more likely to include blinding and a placebo for several reasons. First, industry is less likely to perform pragmatic trials that test an intervention in real-world practice; this absence of real-world conditions makes it more feasible to include double-blinding and a placebo control.

Second, industry entities manufacture not only the drug but also the matching placebo, facilitating easy access. Third, placebos are expensive, and industry-funded trials are typically better resourced than non–industry-funded trials.^7,15^ Fourth, there is often little incentive for a pharmaceutical company to use an available medication as their comparator, as opposed to a placebo; importantly, this can be problematic in cases where the use of an active comparator instead of placebo would have generated more clinically important data.

Another important consideration is the primary outcome itself. We observed that industry-sponsored trials were more likely to utilize surrogate outcomes, which are arguably less clinically relevant to patients and clinicians. And while we did observe that industry-funded trials were smaller than non–industry-funded trials, the absolute difference was modest (i.e., 67 participants).

We found that industry-funded trials are more likely to make their results publicly available on ClinicalTrials.gov. Our results align with a recent study of 4209 trials registered on ClinicalTrials.gov which identified that while two-thirds of the registered trials posted their results, the odds of this occurring were three-fold higher for industry-sponsored trials compared with non–industry sponsored trials.^16^ Similarly, Anderson et al. (2015) found that adherence to legal obligations (outlined by the Food and Drug Administration Amendments Act) for timely reporting of results within 12 months of trial completion was higher among industry-sponsored trials than NIH funded or academic institution funded trials.^17^ This observation might reflect that industry-funded RCTs are more likely to be for new molecules, which legally obligates the investigators to adhere to stricter regulatory reporting.

While reporting compliance appears to be higher among industry-funded trials, it is important to highlight prior literature documenting publication bias among this group. There are instances of industry-funded trials not publishing at all if the trial was negative, or choosing to present positive results while withholding negative results.^18–22^ Consider the previously cited example of Paxil by GlaxoSmithKline, which was prescribed to adolescents for years before it was confirmed to not only be ineffective, but to cause serious side effect including self-injury and suicide.^10^ Or the example of Roche’s Tamiflu (oseltamivir): the initial results from industry-funded trials indicated it was effective in reducing hospitalizations and serious complications from influenza, which resulted in governments stockpiling Tamiflu in advance of potential influenza outbreaks.^23^ However, a Cochrane review subsequently identified that 60% of patient data from the randomized, placebo controlled, phase 3 treatment trials were never published, and that the exclusion of these data significantly changed the findings; in reality, oseltamivir was less effective in reducing complications of influenza than previously stated.^24^

While our study found that trials funded by industry were more likely to have a positive outcome, there are multiple reasons why this might be the case. Publication bias may explain why there was a higher number of positive industry-funded trials; if results aren’t positive, researchers are less likely to pursue publication. Industry-funded trials might be more likely to be positive because such studies focus their resources on drugs and devices that are more likely to work based on earlier phase studies, or involve comparison against placebo rather than an established effective medication. We also found that the concluding statements of trials funded by industry were more likely to have greater positive sentiment than those not funded by industry, which aligns with the greater likelihood of industry-funded trials to have positive outcomes. Our study did not find evidence to either refute or support the claim that industry-sponsored research chooses to withhold negative results.

One limitation of our study is that we lacked data on publication bias because we only included studies that were published. Another limitation is that we focused on three of the highest impact medical journals, and thus our results may not apply to RCTs published in other medical journals. Finally, we lacked data on other aspects of trial design that can directly affect internal validity, including concealment of allocation, number of study sites (e.g., single-centre compared to multi-centre) adherence, statistical analysis (e.g., intention to treat), and loss to follow up.

Taken in sum, our findings suggest that industry-funded RCTs published in high impact journals (i.e., *NEJM, JAMA, Lancet*) are more likely to be blinded, more likely to include a placebo, and more likely to post trial results on ClinicalTrials.gov. Industry funding has been instrumental to major trials with clinical importance, so a culture of systemically undervaluing such trials is concerning. Our findings emphasize the importance of evaluating the quality of an RCT based on its methodological rigour, not its funder type. Our observations also raise questions about how we can empower non–industry-funded research to improve performance in some of these metrics of trial quality. Further research is needed to determine trial characteristics of (a) industry-funded research that is not published and (b) industry-sponsored research that is published in other journals, as the findings from this current study may not be generalizable to these other areas.

## Data Availability

All data produced in the present study are available upon reasonable request to the authors.

## REFERENCES

1. Geoffrey Marshall, Professor J. W. S. Blacklock, Professor C. Cameron, Professor N. B. Capon, Dr. R. Cruickshank, Professor J. H. Gaddum, Dr. F. R. G. Heaf, Professor A. Bradford Hill, Dr. L. E. Houghton, Dr. J. Clifford Hoyle, Professor H. Raistrick, Dr. J. G. Scadding, Professor W. H. Tytler, Professor G. S. Wilson, Dr. P. D’Arcy Hart and the Medical Research Council. STREPTOMYCIN treatment of pulmonary tuberculosis. Br Med J. 1948;2(4582):769–782.

2. Bhatt A. Evolution of clinical research: a history before and beyond james lind. Perspect Clin Res. 2010;1(1):6–10.

3. Kendall JM. Designing a research project: randomised controlled trials and their principles. Emerg Med J. 2003;20(2):164–168.

4. Stolberg HO, Norman G, Trop I. Randomized Controlled Trials. American Journal of Roentgenology. 2004;183(6):1539–1544.

5. Suissa S. Immortal time bias in pharmaco-epidemiology. Am J Epidemiol. 2008;167(4):492–499.

6. Sertkaya A, Wong HH, Jessup A, Beleche T. Key cost drivers of pharmaceutical clinical trials in the United States. Clin Trials. 2016;13(2):117–126.

7. Chopra SS. Industry Funding of Clinical Trials: Benefit or Bias? JAMA. 2003;290(1):113–114.

8. Bodenheimer T. Uneasy Alliance — Clinical Investigators and the Pharmaceutical Industry. N Engl J Med. 2000;342(20):1539–1544.

9. Keller MB, Ryan ND, Strober M, et al. Efficacy of paroxetine in the treatment of adolescent major depression: a randomized, controlled trial. J Am Acad Child Adolesc Psychiatry. 2001;40(7):762–772.

10. Le Noury J, Nardo JM, Healy D, et al. Restoring Study 329: efficacy and harms of paroxetine and imipramine in treatment of major depression in adolescence. BMJ. 2015;351. doi:10.1136/bmj.h4320

11. Kesselheim AS, Robertson CT, Myers JA, et al. A randomized study of how physicians interpret research funding disclosures. N Engl J Med. 2012;367(12):1119–1127.

12. Fralick M, Bartsch E, Darrow JJ, Kesselheim AS. Understanding when real world data can be used to replicate a clinical trial: A cross-sectional study of medications approved in 2011. Pharmacoepidemiol Drug Saf. 2020;(July):1–6.

13. Yang X, Bian J, Hogan WR, Wu Y. Clinical concept extraction using transformers. J Am Med Inform Assoc. 2020;27(12):1935–1942.

14. Devlin J, Chang MW, Lee K, Toutanova K. BERT: Pre-training of Deep Bidirectional Transformers for Language Understanding. In: Proceedings of the 2019 Conference of the North. Association for Computational Linguistics; 2019. doi:10.18653/v1/n19-1423

15. Gelijns AC, Thier SO. Medical innovation and institutional interdependence: rethinking university-industry connections. JAMA. 2002;287(1):72–77.

16. DeVito NJ, Bacon S, Goldacre B. Compliance with legal requirement to report clinical trial results on ClinicalTrials.gov: a cohort study. Lancet. 2020;395(10221):361–369.

17. Anderson ML, Chiswell K, Peterson ED, Tasneem A, Topping J, Califf RM. Compliance with Results Reporting at ClinicalTrials.gov. N Engl J Med. 2015;372(11):1031–1039.

18. Ross JS, Mulvey GK, Hines EM, Nissen SE, Krumholz HM. Trial publication after registration in ClinicalTrials.gov: a cross-sectional analysis. PLoS Med. 2009;6(9):e1000144.

19. Goldacre B. Trial sans error: How pharma-funded research cherry-picks positive results [excerpt]. Sci Am. Published online February 13, 2013. https://www.scientificamerican.com/article/trial-sans-error-how-pharma-funded-research-cherry-picks-positive-results/

20. Djulbegovic B, Lacevic M, Cantor A, et al. The uncertainty principle and industry-sponsored research. Lancet. 2000;356(9230):635–638.

21. Collier R. Is withholding clinical trial results “research misconduct”? CMAJ. 2015;187(10):724–724.

22. Eyding D, Lelgemann M, Grouven U, et al. Reboxetine for acute treatment of major depression: systematic review and meta-analysis of published and unpublished placebo and selective serotonin reuptake inhibitor controlled trials. BMJ. 2010;341:c4737.

23. Gupta YK, Meenu M, Mohan P. The Tamiflu fiasco and lessons learnt. Indian J Pharmacol. 2015;47(1):11–16.

24. Jefferson T, Jones MA, Doshi P, et al. Neuraminidase inhibitors for preventing and treating influenza in adults and children. Cochrane Database Syst Rev. 2014;2014(4):CD008965.

